# Analysis of germline genetic variants that cause hereditary colorectal cancer in bahia

**DOI:** 10.1101/2024.12.06.24318416

**Authors:** Diego S. C. G. Miguel, Maria E. A. V. Alves, Polianna M. Cerqueira, Luíza C. Costa, Rafael N. D. Carvalho, Rodrigo A. Sampaio, David B. O. D. Carvalho, e Iza C. S. Castro

**Affiliations:** Escola Bahiana de Medicina e Saude Publica

## Abstract

**Background:** Colorectal cancer (CRC) is the third most common cancer and the second leading cause of cancer-related deaths worldwide. Approximately 10% of CRC cases are linked to hereditary germline variants. Understanding regional genetic predispositions is crucial for developing personalized medicine strategies.

**Objective:** This study aims to analyze pathogenic germline variants associated with polyposis and non-polyposis syndromes in individuals from Bahia, Brazil.

**Methods:** A cross-sectional, observational study was conducted on 3,100 probands from a private laboratory in Salvador, Bahia, between August 2017 and February 2023. Probands underwent Next Generation Sequencing (NGS) targeting 37 genes. Variants classified as pathogenic (P) or probably pathogenic (PP) in 11 high/moderate penetrance genes were analyzed.

**Results:** Among the 3,100 probands, 97 (3.12%) had P/PP variants. Polyposis syndromes accounted for 47 cases (1.51%), with prevalent variants in MUTYH, APC, and PTEN genes. Non-polyposis (Lynch) syndrome was observed in 50 cases (1.61%), predominantly involving MSH2 and MLH1 genes. Notably, a novel variant, MLH1 c.1127_1130dup, was identified.

**Conclusion:** This study highlights the genetic diversity in CRC predisposition in Bahia, emphasizing the need for targeted regional genetic screening and personalized healthcare strategies. Identifying recurrent pathogenic variants suggests possible shared ancestry among individuals, offering insights for future genetic counseling and public health policies.

## BACKGROUND

Colorectal cancer (CRC) is the third most common cause of cancer and the second most common cause of cancer-related death worldwide (1). Most cases are considered sporadic and are not due to hereditary factors. However, 10% of CRC cancer diagnoses occur in individuals who carry a pathogenic or likely pathogenic germline variant that confers an increased risk of cancer (2). Furthermore, in recent decades there has been a considerable increase in CRC in young adults, reinforcing the need for the identification and diagnosis of hereditary cancer predisposition syndromes (3).

The increase in life expectancy has allowed the expansion of research into familial cancer in medical genetics, leading to the development of diagnostic criteria for specific hereditary syndromes. In this context, describing new pathogenic variants associated with syndromes classified as polyposis or non-polyposis is essential.

### POLYPOSIS SYNDROMES

The main polyposis syndromes that present an increased risk of developing CRC are Familial Adenomatous Polyposis (FAP), which accounts for 1% of all CRC diagnoses, with a prevalence ranging from 1: 6,850 to 1: 31,250 live births (4). Juvenile Polyposis Syndrome has a worldwide prevalence of 1: 100,000 individuals. Peutz-Jeghers Syndrome (PJS), a rare hamartomatous polyposis syndrome, affects 1: 50,000–200,000 people and increases the risk of cancer in the luminal gastrointestinal tract, including the stomach, small intestine, and colon (5). MUTYH-associated polyposis (PAM) is responsible for a smaller proportion of CRCs (6). Hamartomatous syndrome, the main representative of which is Cowden syndrome, has an estimated worldwide prevalence of 1: 200,000 individuals (7).

### NON-POLYPOSIS SYNDROME

The main non-polyposis syndrome is Lynch syndrome, which is characterized by an increased risk of CRC and cancers of the endometrium, ovary, stomach, small intestine, urinary tract, biliary tract, brain, skin, pancreas, and prostate. The prevalence in the world population is estimated at 1:279 (8), occurring in approximately 3% of CRC cases and 10% of cases in probands under 50 years of age (9).

This study aims to describe the pathogenic variants in genes related to polyposis syndromes and Lynch Syndrome identified in Bahia patients.

## METHODS

This is a cross-sectional, observational, analytical, descriptive study.

The study was carried out in a private laboratory in the city of Salvador/BA, of medium size and regional level, which performed the genetic panel on these probands, from August 2017 to February 2023.

The study sample is based on pathogenic genetic variants identified in a germline genetic panel in individuals referred by their attending physicians due to clinical suspicion of Hereditary Cancer Syndrome.

The inclusion criteria are based on individuals with a clinical diagnosis of Cancer Predisposition Syndrome, who underwent germline genetic panel testing, and who agreed to participate in the study. The exclusion criterion is related to previously identified familial mutation.

The sample consisted of genetic data from 3,100 probands sent for the Next Generation Sequencing (NGS) panel, encompassing 37 genes. Pathogenic or likely pathogenic variants found in 11 genes with high or moderate penetrance associated with hereditary polyposis or non-polyposis colorectal cancer were selected.

The database was formulated using the Sophia DDM™ software, which facilitated the collection of variables used in the study without compromising the identification of the individuals participating in the research. The data obtained were analyzed in a private laboratory performing the genetic panel. The variables analyzed were Gene (*APC, MUTYH, STK11, PTEN, SMAD4, BMPR1A, MLH1, MSH2, MSH6, PMS2, EPCAM*), genetic variant (named according to criteria of the Human Genome Variation Society) and variant classification.

**Table 1.** Association of the genes with the syndromes related to CCR.

| Syndromes | Gene |
| --- | --- |
| Classic Familial Adenomatous Polyposis (FAP)<br>Attenuated FAP (AFAP) | *APC* |
| *MUTYH*-Associated Polyposis | *MUTYH* |
| Peutz- Jeghers Syndrome (SPJ) | *STK11* |
| *PTEN*-Associated Hamartomatous syndrome | *PTEN* |
| Juvenile Polyposis Syndrome | *SMAD4* and *BMPR1A* |
| Lynch syndrome | *MLH1*, *MSH2*, *MSH6* and *PMS2* |

Source: Data from the authors

Regarding ethical aspects, the project was submitted to the Comitê de Ética em Pesquisa (CEP), the research ethics committee, of the Escola Bahiana de Medicina e Saúde Pública, and was approved.

## RESULTS

Thus, of the 3,100 probands, 97 (3.12%) individuals presented pathogenic or probably pathogenic (P/PP) germline genetic variants in some of the 11 genes. Of this group, 47 (1.51%) individuals had P/PP variants associated with Polyposis Syndromes, while 50 (1.61%) presented P/PP variants associated with Lynch Syndrome.

In the 97 individuals, 44 P/PP variants were found in the genes studied, 23 variants associated with Lynch Syndrome and 21 variants associated with polyposis syndromes.

### Polyposis Syndromes

In decreasing order of prevalence, *MUTYH* (25 probands), *APC* (12 probands), *PTEN* (9 probands) and *STK11* (1 proband). The most frequent pathogenic variant was associated with the *PTEN* gene c. 802-2A>G, found in 8 probands. It is worth noting that all cases with variants in the *MUTYH* gene were in simple heterozygosity (Table 2). No P/PP variants were found in the *BMPR1A* and *SMAD4* genes.

**Table 2.** P/PP variants found in genes associated with Polyposis Syndromes Bahia/2017-2023.

| Gene | cDNA | Protein | Probands |
| --- | --- | --- | --- |
| APC/ NM_000038 | c.637C>T | p.(Arg213*) | 1 |
|  | c.1100_1101del | p.(Ser367Cysfs*10) | 1 |
|  | c.4638_4642del | p.(Asn1546Lysfs*11) | 1 |
|  | c.1972_1975del | p.(Glu658Thrfs*11) | 1 |
|  | c.2247G>T | p.Leu749Phe | 7 |
|  | c.3747C>A | p.(Cys1249*) | 1 |
| MUTYH/ NM_001048174 | c.452A>G | p.(Tyr151Cys) | 6 |
|  | c.1103G>A | p.(Gly368Asp) | 4 |
|  | c.691C>T | p.Arg231Cys | 1 |
|  | c.1143_1144dup | p.(Glu382Glyfs*43) | 1 |
|  | c.247C>T | p.(Arg83*) | 1 |
|  | c.1063del | p.(Ala357Profs*23) | 2 |
|  | c.763A>G | p.(Met255Val) | 1 |
|  | c.692G>A | p.(Arg231His) | 3 |
|  | c.637C>T | p.(Arg213Trp) | 1 |
|  | c.463-2A>C | - | 1 |
|  | c.694G>T | p.(Val232Phe) | 3 |
|  | c.463-2A>G | - | 1 |
| PTEN/ NM_000314 | c.1003C>T | p.Arg335* | 1 |
|  | c.802-2A>G | - | 8 |
| STK11/ NM_000455 | c.816C>A | p.(Tyr272*) | 1 |

Source: Data from the authors

**Graphic 1:**
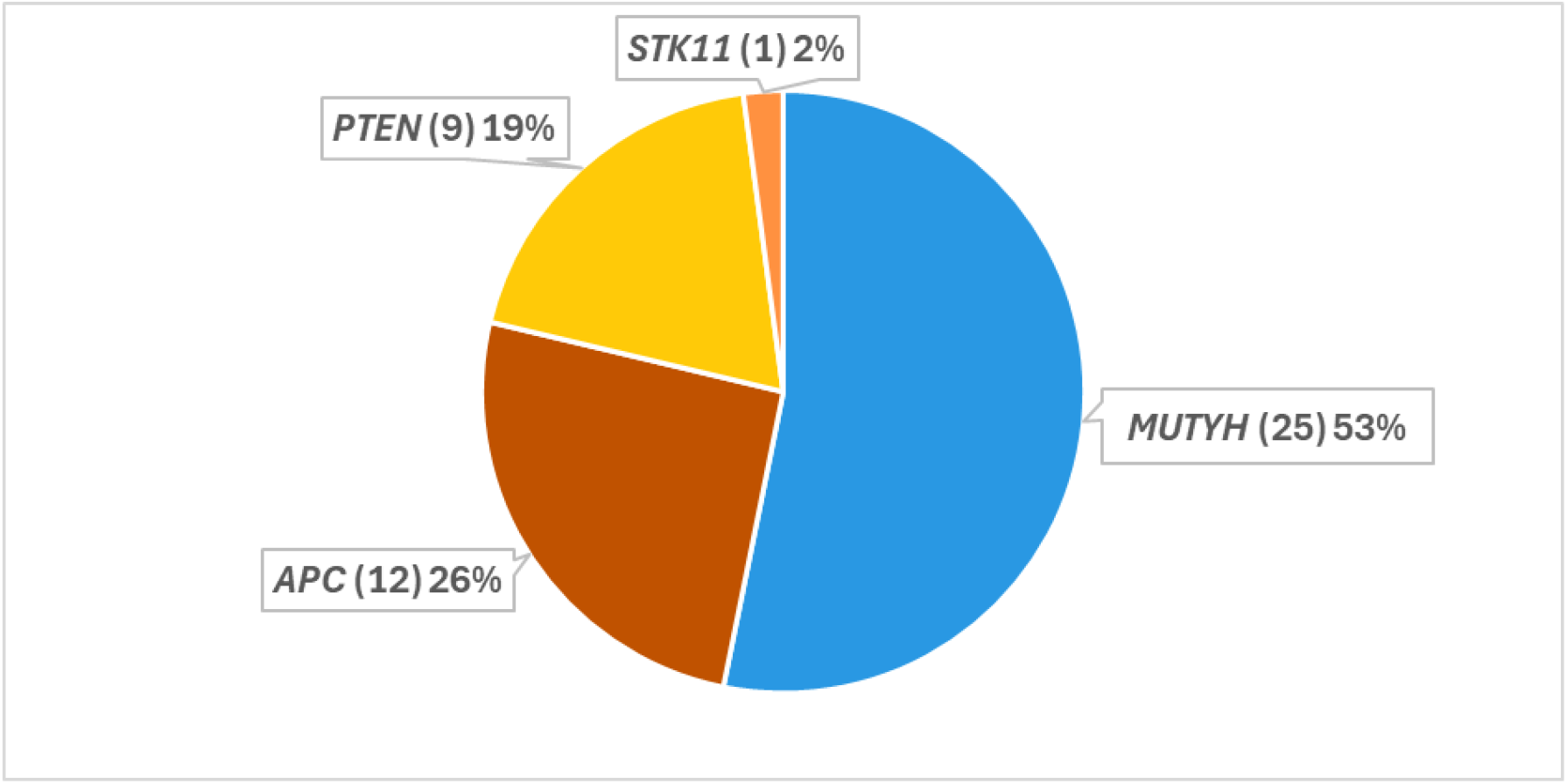
Absolute and relative number of individuals with P/PP variants in genes associated with Polyposis Syndromes Bahia/2017-2023. Source: Data from the authors

### Nonpolyposis Syndrome (HNPCC - Lynch)

In decreasing order of prevalence, *MSH2* (27 probands), *MLH1* (12 probands), *PMS2* (9 probands) and *MSH6* (2 probands). The most frequent pathogenic variant was c.187dupG, in the *MSH2* gene, present in 13 probands. In addition, a new variant, not yet described in the literature, c.1127_1130dup, associated with the *MLH1* gene present in one proband was identified (Table 3).

**Table 3.** P/PP variants found in genes associated with Lynch Syndrome Bahia/2017-2023.

| Gene | cDNA | Protein | Probands |
| --- | --- | --- | --- |
| *MLH1*/ NM_000249 | c.793C>T | p.(Arg265Cys) | 1 |
|  | c.1772_1775del | p.(Asp591Valfs*24) | 2 |
|  | c.350C>T | p.Thr117Met | 1 |
|  | c.588+5G>C | - | 7 |
|  | c.1127_1130dup | p.(Val378*) | 1 |
| *MSH2*/ NM_000251 | c.187dupG | p.(Val63Glyfs*19) | 13 |
|  | c.1705_1706delG<br>A | p.Glu569Ilefs*2 | 1 |
|  | c.2131C>T | p.(Arg711*) | 4 |
|  | c.388_389del | p.(Gln130Valfs*2) | 2 |
|  | c.1447G>T | p.(Glu483*) | 1 |
|  | c.1444A>T | p.(Arg482*) | 1 |
|  | c.1984C>T | p.(Gln662*) | 1 |
|  | c.2005+2delT | - | 1 |
|  | c.2006-1G>C | - | 1 |
|  | c.645+1G>T | - | 1 |
|  | c.1193del | p.(Ala398Glufs*14) | 1 |
| *PMS2*/ NM_000535 | c.1687C>T | p.(Arg563*) | 1 |
|  | c.1731_1732delin<br>sAGT | p.(Arg578Valfs*3) | 1 |
|  | c.631C>T | p.(Arg211*) | 1 |
|  | c.2192_2196del | p.(Leu731Cysfs*3) | 1 |
|  | c.137G>T | p.Ser46Ile | 1 |
|  | c.1A>G | p.(Met1Val | 4 |
| *MSH6*/ NM_000179 | c.1519dupA | p.(Arg507Lysfs*9) | 2 |

Source: Data from the authors

**Graphic 2:**
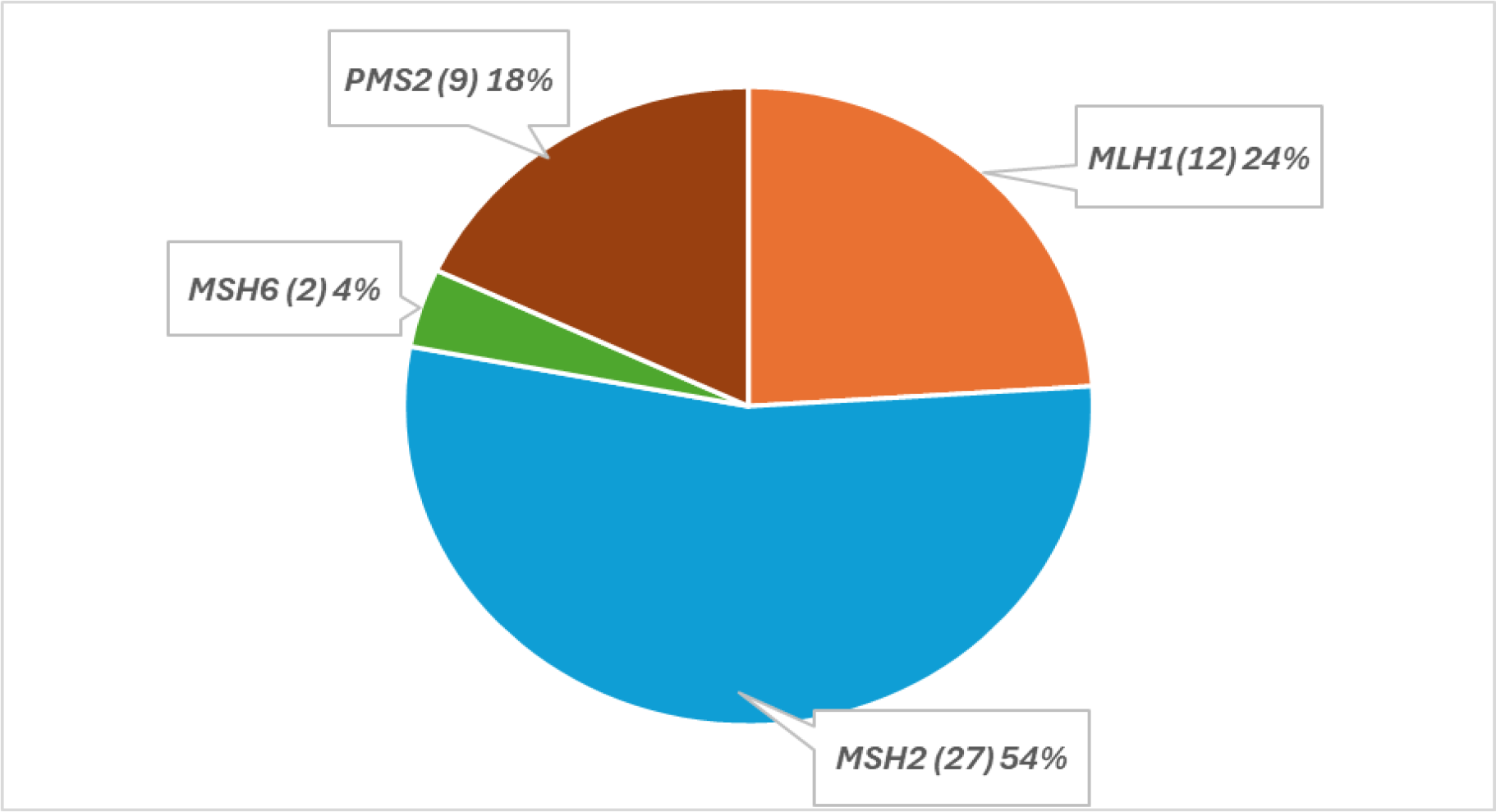
Absolute and relative number of individuals with P/PP variants in genes associated with Lynch Syndrome Bahia/2017-2023Source Source: Data from the authors

## DISCUSSION

Regarding Polyposis Syndromes, according to the literature, the highest prevalence of pathogenic variants occurs in the *APC* gene, with a prevalence in the world population of 1:6,850 to 1:31,250 individuals (4). In the sample analyzed in this study, a higher prevalence of pathogenic variants was observed in the *MUTYH* gene, totaling 25 individuals. This divergence in the findings can probably be attributed to the fact that most probands with Familial Adenomatous Polyposis are clinically diagnosed and are not referred by their attending physicians for genetic testing. In addition, the most prevalent variant occurred in the *PTEN* gene, c.802-2A>G, with 8 probands, and there were no cases of pathogenic variants in the *SMAD4* and *BMPR1A* genes.

Regarding Lynch Syndrome, according to the literature, most pathogenic variants occur in the *PMS2* gene, with a prevalence in the world population of 1:714 individuals (8). However, screening for germline variants in *PMS2* is technically challenging due to homology with its pseudogene *PMS2CL*. Sequences of *PMS2* and *PMS2CL* are so similar that NGS of short fragments identify the presence of a variant but fail to disambiguate whether its origin is the gene or the pseudogene, which may result in the identification of a pathogenic variant that is not real (10). In the sample analyzed in this study, a higher prevalence was observed in the *MSH2* gene, in 27 probands. The most prevalent variant was c.187dupG also in this gene, present in 13 probands. There were no cases of pathogenic variants in the *EPCAM* gene. Furthermore, a variant, not yet described in the literature, c.1127_1130dup in the *MLH1* gene, present in one patient, was found.

The individuals analyzed in this study were referred for genetic panel testing for reasons that do not qualify as family screening for Cancer Predisposition Syndrome. However, the finding of the same pathogenic variant in a considerable number of probands, as occurred in the variants c.802-2A>G (*PTEN*), c.2247G>T (*APC)*, c.452A>G (*MUTYH*), c.187dupG (*MSH2*), and c.588+5G>C (*MLH1*), raises the hypothesis of a possible unknown common ancestry among these individuals or that they are individuals from the same family referred for different reasons, justifying this finding given that these are rare syndromes.

## CONCLUSION

Therefore, this study is extremely relevant in the production of regional data on the epidemiological profile, assisting in the development of more specific and assertive public policies for this population. Thus, promoting the practice of evidence-based personalized medicine.

## Data Availability

All data produced in the present study are available upon reasonable request to the authors

## DECLARATIONS

### Ethics, Consent to Participate, and Consent to Publish

This study was conducted in accordance with the principles of the Declaration of Helsinki and relevant guidelines and regulations. All methods were performed in compliance with these standards and approved by the Comitê de Ética em Pesquisa da Escola Bahiana de Medicina e Saúde Pública (approval number: CAAE 75073223.6.0000.5544). Informed consent was obtained from all participants and/or their legal guardians. Additionally, written consent for the publication of anonymized data was obtained.

## Funding

This research did not receive any specific grant from funding agencies in the public, commercial, or not-for-profit sectors. The costs associated with the study were covered by the authors.

## Competing Interests

The authors declare no relevant financial or non-financial competing interests.

## Author Contributions

All authors contributed to the conception and design of the study. Material preparation and data collection were performed by Luíza Cajado Costa, Maria Eduarda Amorim Vieira Alves, Polianna Massa Cerqueira, David Bastos Oliveira Diniz Carvalho, Rafael Nascimento de Carvalho, and Rodrigo Amazonas Sampaio. Data analysis and manuscript writing were conducted by Diego Santana Chaves Geraldo Miguel, Maria Eduarda Amorim Vieira Alves and Polianna Massa Cerqueira. Iza Cristina Sales de Castro, Rejane Hughes Castro, and Diego Santana Chaves Geraldo Miguel reviewed and provided comments on earlier versions of the manuscript. All authors read and approved the final manuscript.

## Consent for Publication

Consent for the publication of data was obtained from all participants.

## Acknowledgments

We would like to express our sincere gratitude to Professor Maria Betânia Toralles for providing access to the essential data that made this study possible and for her invaluable encouragement in promoting scientific research. Her support and guidance were fundamental to the successful completion of this work.

## Notes

### Competing Interest Statement

The authors have declared no competing interest.

### Funding Statement

This study did not receive any funding

### Author Declarations

Ethics committee/Escola Bahiana de Medicina e Saude Publica gave ethical approval for this work (approval number: CAAE 75073223.6.0000.5544).

